# Wastewater surveillance for SARS-CoV-2 on college campuses: Initial efforts, lessons learned and research needs

**DOI:** 10.1101/2021.02.01.21250952

**Authors:** Sasha Harris-Lovett, Kara Nelson, Paloma Beamer, Heather N. Bischel, Aaron Bivins, Andrea Bruder, Caitlyn Butler, Todd D. Camenisch, Susan K. De Long, Smruthi Karthikeyan, David A. Larsen, Katherine Meierdiercks, Paula Mouser, Sheree Pagsuyoin, Sarah Prasek, Tyler S. Radniecki, Jeffrey L. Ram, D. Keith Roper, Hannah Safford, Samendra P. Sherchan, William Shuster, Thibault Stalder, Robert T. Wheeler, Katrina Smith Korfmacher

**Author notes:** **Corresponding Author:** Sasha Harris-Lovett, Ph.D., 410 O’Brien Hall, University of California Berkeley, Berkeley, CA 94720. The authors declare they have no actual or potential competing financial interests.

## Abstract

**Background:** Wastewater surveillance for SARS-CoV-2 is an emerging approach to help identify the risk of a COVID-19 outbreak. This tool can contribute to public health surveillance at both community (wastewater treatment system) and institutional (e.g., colleges, prisons, nursing homes) scales.

**Objectives:** This research aims to understand the successes, challenges, and lessons learned from initial wastewater surveillance efforts at colleges and university systems to inform future research, development and implementation.

**Methods:** This paper presents the experiences of 25 college and university systems in the United States that monitored campus wastewater for SARS-CoV-2 during the fall 2020 academic period. We describe the broad range of approaches, findings, resource needs, and lessons learned from these initial efforts. These institutions range in size, social and political geographies, and include both public and private institutions.

**Discussion:** Our analysis suggests that wastewater monitoring at colleges requires consideration of information needs, local sewage infrastructure, resources for sampling and analysis, college and community dynamics, approaches to interpretation and communication of results, and follow-up actions. Most colleges reported that a learning process of experimentation, evaluation, and adaptation was key to progress. This process requires ongoing collaboration among diverse stakeholders including decision-makers, researchers, faculty, facilities staff, students, and community members.

## Introduction

Since the spring of 2020, many colleges have pursued wastewater monitoring for SARS-CoV-2, the virus that causes COVID-19, as part of a multi-pronged approach to controlling COVID-19 transmission on campus. In August 2020, the University of Arizona made headlines by announcing that it had detected RNA from SARS-CoV-2 in the wastewater from a student dormitory (Peiser 2020). Subsequent testing of dormitory residents identified two asymptomatic infected students, who were transferred to an isolation facility, potentially preventing an outbreak of COVID-19 on campus (Betancourt et al. 2020). As colleges across the country considered their options for reducing transmission of COVID-19, the University of Arizona story piqued interest in wastewater monitoring as a promising tool. By the authors’ count, news media in the United States published nearly 200 articles on wastewater monitoring on college campuses in September 2020 alone (for this paper, we use the term “colleges” to describe institutions of higher education, including colleges, universities, and university systems spanning multiple campuses).

As of January 2021, more than 210 colleges around the world had begun monitoring wastewater for SARS-CoV-2 (University of California Merced 2021), and many more are considering launching similar efforts. Our synthetic, comparative study found that institutions’ approaches to wastewater monitoring vary by where, how, and how often they sample, their analytical and reporting protocols, and the use of their findings in decision-making. The lessons learned from these emerging experiences can inform other colleges, institutions (e.g. nursing homes, prisons, private industries), and communities seeking to manage COVID-19.

This paper synthesizes the experiences of 25 colleges that monitored campus wastewater for SARS-CoV-2 during fall 2020. It describes the broad range of approaches, resource needs, and lessons learned from these initial efforts. These experiences provide early insights into varied approaches, decision-support potential, and research needs related to wastewater surveillance by colleges. Based on these reported experiences, we developed a process-oriented framework for design of wastewater surveillance at colleges. This framework provides a structure for the collaborative learning process needed to successfully implement, evaluate, and adapt wastewater surveillance programs.

## Background

Wastewater-based epidemiology (WBE) has long been used to inform public health decisions about infectious disease, most prominently in the global effort to monitor elimination of polioviruses (Asghar et al. 2014). Similar to polioviruses, SARS-CoV-2 RNA is shed by many infected individuals in fecal matter (Wölfel et al. 2020) and is relatively stable in wastewater (Ahmed, Bertsch, et al. 2020). Soon after the start of the pandemic, researchers around the world began developing methodologies to detect SARS-CoV-2 RNA in sewage (Ahmed, Angel, et al. 2020; Medema, Heijnen, et al. 2020; Bivins et al. 2020; Gonzalez et al. 2020; Sherchan et al. 2020). Methods generally involve concentration of viral particles in wastewater, and molecular biology assays that measure SARS-CoV-2 RNA (Philo et al. 2021).

Researchers continue to refine sample collection and data analysis with the goal of providing a real-time quantitative indicator of prevalence, increase, and geographic reach of COVID-19 within a population (Ahmed, Bivins, et al. 2020; Farkas et al. 2020; Graham et al. 2021; Peccia et al. 2020). Meanwhile, several U.S. cities have begun monitoring for SARS-CoV-2 in sewage at municipal wastewater treatment plants (Stadler et al. 2020; Gonzalez et al. 2020; Wu et al. 2020; Sherchan et al. 2020). In some cases, these data are made public on online dashboards, and accompanied by guidance for public health messaging (e.g., Ohio Department of Health 2021; Oregon Health Authority 2021). To support these efforts, the Centers for Disease Control and Prevention (CDC) has established a National Wastewater Surveillance System (CDC 2020).

Monitoring wastewater for SARS-CoV-2 is a useful complement to clinical surveillance for COVID-19 (Bivins et al. 2020; Farkas et al. 2020; Larsen and Wigginton 2020). Wastewater monitoring has particular value when clinical testing is limited (Peccia et al. 2020). In addition, the SARS-CoV-2 wastewater signal may be a leading indicator that precedes trends in confirmed cases (Wu et al. 2020; Larsen and Wigginton 2020; Randazzo, Truchado, et al. 2020). This early warning from wastewater may occur because wastewater monitoring detects both pre-symptomatic and asymptomatic SARS-CoV-2 infections (Buitrago-Garcia et al. 2020; Wang et al. 2020). Wastewater monitoring also provides cost-effective infection information about a large population (Randazzo, Cuevas-Ferrando, et al. 2020; Hart and Halden 2020). If used to target allocation of pandemic-response resources, this approach could help offset the inequitable impacts of the pandemic.

Because of these advantages, many researchers, government agencies, and communities have promoted wastewater monitoring as an important component of pandemic response (Bivins et al. 2020; Farkas et al. 2020; Medema, Been, et al. 2020; Xagoraraki 2020). Wastewater results could be used to alert communities to increased COVID-19 prevalence and track spread, guide individual behavioral choices, target public health messaging, allocate testing resources, inform infection control policies (e.g. limiting size of gatherings, building openings, and school modalities), and evaluate the success of such interventions (Daughton 2020; Farkas et al. 2020; Polo et al. 2020; Hassard et al. 2021). Although some have raised concerns about privacy, stigma, and potential negative repercussions of sharing these data (Joh 2020), the community-wide, non-individualized nature of the technology mitigates potential legal and ethical issues of using wastewater monitoring for public health purposes (Gable, Ram, and Ram 2020).

As wastewater monitoring for public health surveillance has gained traction in the United States, many colleges have initiated and implemented wastewater-monitoring programs to address an urgent need to monitor for potential infections on campus. Several professional networks have emerged to support co-learning, including a website (“CoSeS: Communicating Sewage Surveillance for COVID-19” 2021), a Slack channel, and an NSF-funded Research Coordination Network (Research Coordination Network: Wastewater Surveillance of SARS-CoV-2 2020). The National Academies of Sciences Engineering and Medicine undertook a “rapid expert assessment” of COVID-19 surveillance efforts at colleges, many of which integrated wastewater monitoring (National Academies 2020). However, there has not been a systematic effort to review the experiences of colleges’ pioneering efforts and to synthesize lessons learned. This paper represents a first step: to collect insights from colleges on wastewater monitoring for SARS-CoV-2 in order to inform future research and action.

## Methods

Case studies were solicited through email lists, Slack channels, and informal networks among practitioners conducting wastewater monitoring at colleges. Respondents – largely faculty and staff involved in these efforts – were asked to self-report descriptions of their institution’s history, practice, and use of wastewater monitoring for SARS-CoV-2 on campus via a shared database. All participants were given the opportunity to check the accuracy of their college’s portrayal in the paper and to clarify any ambiguous responses.

Open-ended interviews were conducted with a subset of respondents from 10 colleges with diverse experiences to elicit in-depth lessons learned about wastewater monitoring on their campuses. Interview protocols were approved by the Institutional Review Boards at the University of California Berkeley and the University of Rochester. Interview notes were separately coded for common themes, observations, and recommendations by Harris-Lovett and Korfmacher, drawing from both the ENTREQ and COREQ protocols for conducting and reporting qualitative research (Tong et al. 2012; Tong, Sainsbury, and Craig 2007). Differences in coding were reconciled through discussion or follow-up with interviewees. Each case study contributor was invited to be a co-author or named contributor.

Details of monitoring programs at participating colleges were corroborated where possible using publicly available websites and/or media reports. The size, residential nature, and location of each institution was similarly confirmed. Distinction was made between private and state institutions, as each is accountable to a different set of stakeholders, regulations, levels of external decision-making, resource constraints, and ability to compel student behavior (e.g. testing requirements). These multiple sources of information were integrated into the case study analyses.

## Results

### Case study institutions

Twenty-five colleges and universities from 16 states in the U.S. provided information about their wastewater monitoring programs (Table 1, Figure 1). Most respondents represented a single campus, although respondents from three state university systems (Maine, Oregon State, and Utah State) represented two or more campus locations (Table 1). The case study institutions represent rural, suburban, and urban settings within socially and politically diverse geographies. Approximately two-thirds of participating institutions are public; the remainder are private. Student populations of the campuses/systems ranged from approximately 2,000 to 50,000.

**Table 1:** Characteristics of case study colleges

| College Name | Location City | State | Total 2019 Enrollment | Public/private |
| --- | --- | --- | --- | --- |
| Clemson University | Clemson | SC | 25,822 | public |
| Colorado College | Colorado Springs | CO | 2,270 | private |
| Colorado State University | Fort Collins | CO | 33,996 | public |
| Hope College | Holland | MI | 3,060 | private |
| Oregon State University (system) | Multiple | OR | 28,886 | public |
| St. John Fisher College | Rochester | NY | 3,647 | private |
| Siena College | Loudonville | NY | 3,226 | private |
| SUNY Morrisville | Morrisville | NY | 3,000 | public |
| SUNY Oneonta | Oneonta | NY | 6,733 | public |
| Syracuse University | Syracuse | NY | 22,850 | private |
| Tulane University | New Orleans | LA | 14,602 | private |
| University of Arizona | Tucson | AZ | 45,918 | public |
| University of California Berkeley | Berkeley | CA | 42,347 | public |
| University of California Davis | Davis | CA | 39,629 | public |
| University California San Diego | San Diego | CA | 38,396 | public |
| University of Connecticut | Mansfield | CT | 32,333 | public |
| University of Georgia | Athens | GA | 38,920 | public |
| University of Idaho | Moscow | ID | 10,791 | public |
| University of Maine (system) | Multiple | ME | 35,337 | public |
| University of Massachusetts Amherst | Amherst | MA | 49,617 | public |
| University of Massachusetts Lowell | Lowell | MA | 18,338 | public |
| University of New Hampshire | Durham | NH | 14,509 | public |
| University of Notre Dame | Notre Dame | IN | 11,836 | private |
| Utah State University | Multiple | UT | 27,691 | public |
| Wayne State University | Detroit | MI | 26,251 | public |

**Figure 1.**
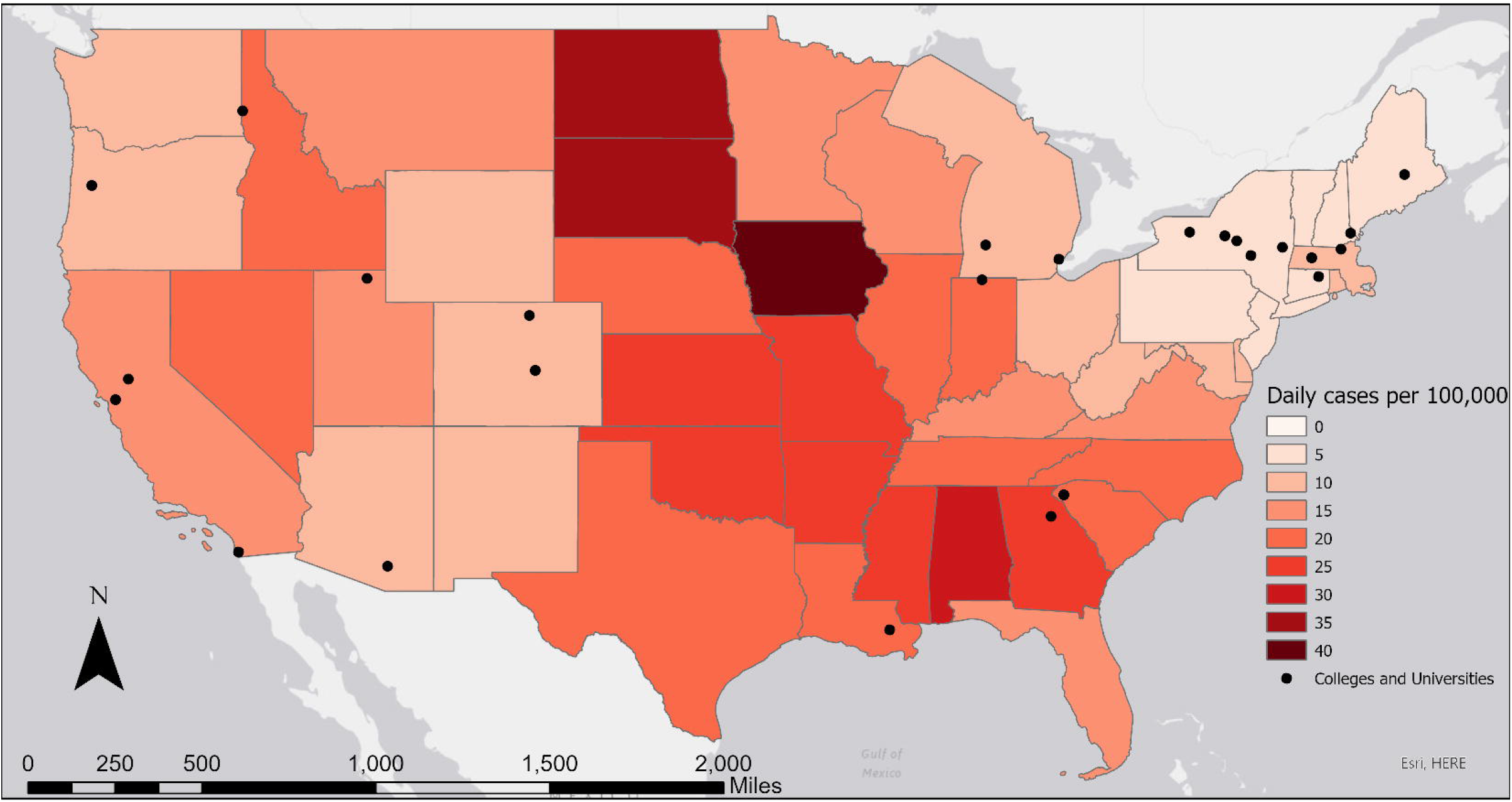
Map showing location of case study colleges and average daily number of COVID-19 cases per 100,000 population during the last week of August 2020, by state. Data from Centers for Disease Control and Prevention, 2020 (CDC (Centers for Disease Control and Prevention) 2020a). Note: A single dot in Utah, Oregon, and Maine represents a system of more than one university in each state that work together on wastewater surveillance.

Some of the larger campuses have a dedicated wastewater treatment plant, while other campuses are served by the wastewater infrastructure of the surrounding community. Larger universities in urban locations generally have a smaller proportion of residential (e.g., on-campus) students and a correspondingly larger proportion of students living off-campus in the surrounding community. The smaller colleges had close to their normal, pre-pandemic number of students living on and around campus. The majority of participating colleges offered some mix of virtual and in-person course options in the fall 2020 academic period. Even those that offered entirely remote instruction had some students living in campus housing.

The colleges started the fall 2020 academic period with considerable variation in COVID-19 case rates in the surrounding area, ranging from 1.5 (New Hampshire) to 20.4 (Georgia) daily new cases per 100,000 population (Figure 1). In many places with low COVID-19 rates, local communities voiced concerns about students carrying the virus from other states and countries. Thus, colleges designed their surveillance systems under very different community conditions, with significant implications for local public health and campus/community relationships.

### Origins and organization of wastewater monitoring on campus

In many cases, campus researchers seeking to address urgent pandemic-related needs initiated college wastewater monitoring efforts. More than half of the wastewater monitoring programs were started by faculty from engineering disciplines, several in collaboration with biological scientists. Other programs were initiated by faculty in other disciplines (including math, environmental health and epidemiology), by facilities staff, college administrators, or county officials. Regardless of who initiated the program, nearly all reported that a multidisciplinary team of faculty, facilities staff, and student health professionals collaborated to sustain the effort. Many of the faculty involved had longstanding research programs involving pathogens in wastewater and several had engaged in broader wastewater monitoring efforts for SARS-CoV-2 in their region before applying this approach to their colleges. Other respondents, however, pivoted from their previous research to adapt their expertise to wastewater monitoring. All of the respondents noted that their wastewater monitoring efforts interfaced with a range of stakeholders, including college administrators, students, researchers, facilities staff, local public health officials, and/or the surrounding community.

Around half of the colleges started sampling wastewater in August in preparation for the arrival of students. A quarter began sampling earlier (as early as May) as part of methodology development; the remainder did not initiate sampling until mid-fall. Thus, only a small number of the colleges had experience with data from occupied dorms going into the fall semester, but many were able to capture baseline data prior to student move-in.

At a time when most colleges experienced financial challenges, obtaining funding for these efforts was a frequent challenge. Around half of respondents noted their university administrations funded wastewater monitoring efforts. Several participants noted that their administrations “basically wrote a blank check,” acknowledging that optimal surveillance was essential to keeping the campus open, while others cited pressure to control costs. Other funding mechanisms included support from local or state government, federal CARES Act relief funding for coronavirus surveillance, research grant funds, and philanthropic gifts.

### Description of wastewater monitoring approaches

The colleges’ approaches varied with respect to how wastewater samples were collected, sampling locations, how often samples were taken, laboratory analysis, and how results were reported and used (Table 2). These activities were carried out by different groups of faculty, staff, contractors, students and administrators on different campuses. Many respondents reported that their approaches evolved over time as they developed expertise, acquired additional resources, and scaled up their efforts.

**Table 2.**
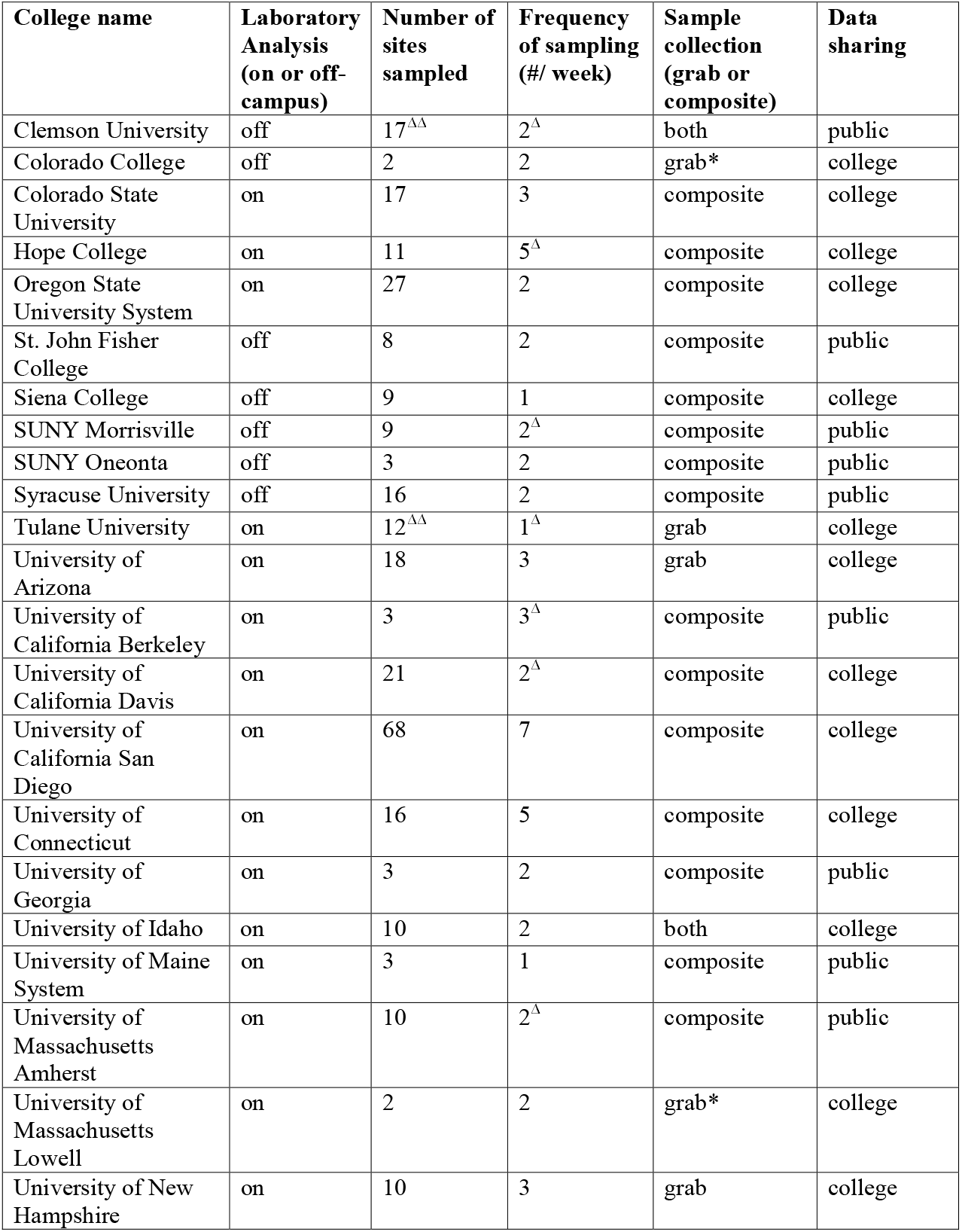
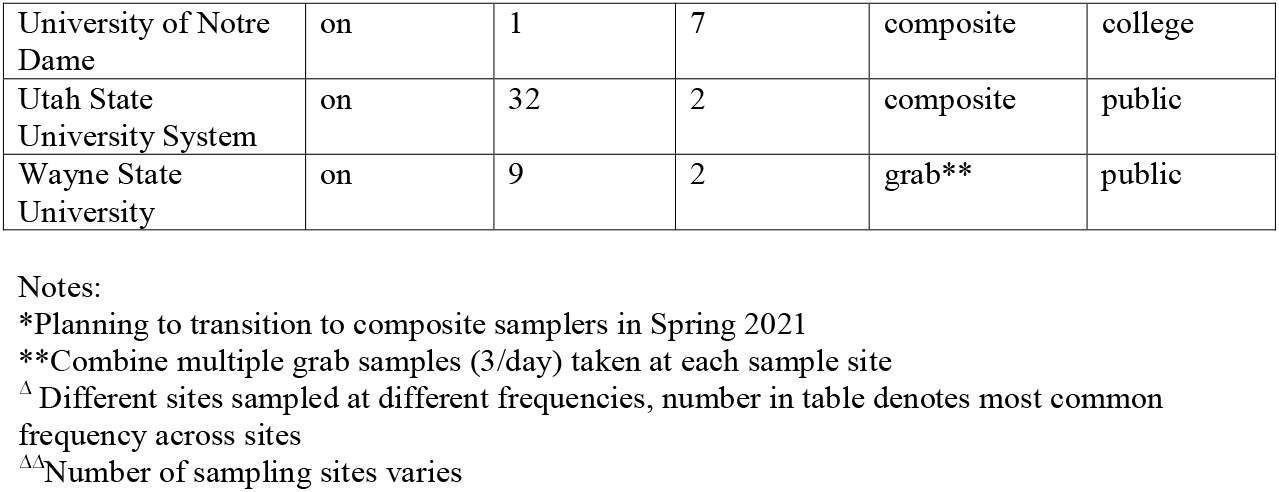
Characteristics of college wastewater monitoring programs

In theory, wastewater samples can be collected from any accessible point in a sewer system, but some are logistically simpler than others. The majority of colleges in this study collected samples from sewer manholes. Several collected wastewater from pipes or sewer cleanouts in dormitories, which can involve significant plumbing alterations. Participants expressed more problems with autosampler clogging (toilet paper, large proportion of solids under low-flow conditions) at smaller-diameter pipes and building cleanouts relative to other sample collection points in their system. Around one-quarter of the colleges took samples of the influent to the municipal wastewater treatment plants serving their campuses. Many of the colleges sampled at multiple locations with differently sized sewer drainages (e.g., dorm, main sewer lines, and wastewater treatment plant). Decisions about sampling location often reflected complex tradeoffs between costs, logistical constraints (e.g. physical access), and ability to associate individual sampling locations with specific student residences.

Wastewater samples can be collected as one-time “grab” samples, as passive samples using absorbent swabs (Liu et al. 2020), or as composite samples. Automated composite samplers collect wastewater aliquots periodically over a 24-hour period to provide a more representative sample of the sewage (Ahmed et al. 2021). Grab samples are normally taken during peak (morning and evening) sewer flows (Curtis et al. 2020). Composite samplers cost between $3,000 to $5,000. Passive samplers are inexpensive, but less is known about their sensitivity for detecting SARS-CoV-2 compared to composite samplers (Liu et al. 2020). Several colleges that could not purchase composite samplers due to cost or supply shortages constructed their own (Kilaru, Larsen, and Monk 2020).

Colleges used student workers, existing staff, or private contractors to collect samples and deliver or ship them to laboratories for analysis. Retrieving samples from collection points took from 15 minutes to 3 hours per sample (not including the 24 hours over which composite samples are collected) depending on the physical layout of sampling locations and equipment.

The number of sampling locations varied from one to more than fifty per campus. Two-thirds of the colleges with normal (i.e. non-pandemic) enrollment of over 10,000 had 10 or more sampling locations. The population size represented by a single wastewater sample ranged from a single dorm to the entire campus community. For campuses that used wastewater monitoring to guide targeted individual testing of all residents in a building, the reported number of students per sampling location ranged from 50 to 800. 28% reported sampling at three or fewer sites. Several sampled only at a local wastewater treatment plant or identified a single manhole in a sewer line collecting most of the flow from the campus. Several respondents noted they had increased their number of sampling locations over time (or planned to do so in the future) to reduce the number of students who would be individually tested as a result of a “hot” wastewater sample.

Colleges reported a range of sampling frequencies, from daily to weekly. Of those that reported sampling once per week, most noted that they are still in the process of developing their surveillance system and planned to increase sampling frequency in the future. Nearly three-quarters (72%) reported taking samples at most locations two or three times per week. 16% responded that some or all of their sites are sampled daily (5-7 times/week). Several noted that samples were taken with different frequencies at different locations, and that sample collection frequency varied over time. For example, when the virus was detected in the wastewater of a specific dorm, they might increase sampling frequency at that location.

The majority of colleges analyzed their samples in on-campus laboratories; 28% used an off-campus commercial laboratory. Of those that analyzed their own samples, the vast majority of them (88%) relied in part or wholly on students (both undergraduates and graduates), postdoctoral fellows, and faculty for wastewater sample analysis. All of the respondents used quantitative analysis methods, either real-time reverse transcriptase quantitative polymerase chain reaction (RT-qPCR) (68%), reverse transcriptase digital droplet polymerase chain reaction (RT-ddPCR) (20%), or both technologies (12%) to identify the number of copies of RNA per mL of wastewater. Colleges employed a range of viral concentration and RNA extraction methods depending on factors including expertise, availability, cost, and speed. A technical comparison of the different laboratory methods each college has been reported elsewhere (Pecson et al. 2021).

Program costs varied based on the number of sampling sites, number of samples analyzed per week, costs per sample, and setup costs. Initial capital investments in equipment and staffing (e.g. to collect and process samples, etc.) ranged from $1,000 (using only existing equipment and facilities, with in-kind support from faculty and students to collect and analyze samples) to over $500,000 (purchases of equipment, hiring new staff, and renting facilities for laboratory space). Costs per sample are not readily comparable across institutions with different models of accounting for labor, overhead and supplies costs. However, those that contracted with private off-campus labs reported analysis fees ranging from $200 to $450 per sample. For those that conducted their own analyses, costs from $20 to $400 per sample were reported (exclusive of labor). Laboratory processing times ranged from 5 hours (for an on-campus lab using ddPCR) to 15 hours (depending on sample turbidity, using qPCR). Those using off-site labs generally used refrigerated shipping services, which added to the per-sample cost.

### Reporting and use of wastewater monitoring results in campus decision-making

All of the respondents reported sharing wastewater monitoring results with campus decision makers. A subset also shared their results with local government (e.g., wastewater agency staff; local, regional, or state health department). Over a quarter of the colleges publicly shared wastewater results via text message or email to residents of affected dorms, in whole-campus email announcements, or by integrating wastewater results into their campus COVID-19 surveillance dashboard.

The colleges communicated using diverse approaches, included establishing categorical thresholds for “Levels” of SARS-CoV-2 RNA in wastewater (e.g. “low,” “medium,” or “high”), providing absolute data (e.g. concentrations of RNA detected), reporting trends for each sampling site (e.g. increasing, decreasing, or stable), or simply noting presence/absence of SARS-CoV-2. Most reports included a summary of the wastewater surveillance process, uncertainties involved, and implications for local public health risks. Some of the colleges included follow-up actions in these communications (e.g. testing of dorm residents, recommending hand-washing and social distancing), whereas others simply reported the results. Some colleges did not regularly report results, but rather integrated wastewater data into messaging as relevant to changes in college policies, such as reducing allowed gathering sizes or moving to remote instruction.

Regardless of how, when, and to whom wastewater results were communicated, nearly all colleges integrated wastewater data into their college’s overall COVID-19 surveillance and response system. Several noted this integration is still a work in progress. Two-thirds reported that a key function of their wastewater monitoring was to target clinical testing (either pooled or individual diagnostic testing, including saliva, nasal swab, or nasal-pharyngeal swab) to students living in residences with elevated SARS-CoV-2 RNA in their wastewater. Several noted that targeting individual testing in response to a wastewater signal was a less costly approach than frequent surveillance testing of all students to identify asymptomatic or pre-symptomatic cases. Even where regular clinical surveillance testing was taking place, wastewater results were helpful in providing early warning of infected individuals in dorms and requiring students to quarantine until tested (CNN (Cable News Network) 2020). In several cases, wastewater results also helped detect risks from untested individuals, including visitors or staff. One college reported using wastewater data to evaluate the effectiveness of university interventions such as email alerts recommending individual testing or reducing gathering size limits. At several colleges, wastewater results corroborated trends in individual test results and gave campus decision makers “more confidence” as they weighed restrictive measures like pausing in-person classes.

Respondents noted that the role of wastewater monitoring results at colleges may change over time. For example, several colleges found that wastewater results were most straightforward in the “maintenance phase” after students were tested post-arrival on campus and before case rates rose significantly. Once a significant number of infected students return to their dorms after isolation, they may continue to shed the virus (Wang et al. 2020), complicating interpretation of wastewater results. Wastewater data is also expected to be highly useful for tracking possible outbreaks after the colleges’ populations begin to get vaccinated and institutions reduce individual testing (Smith, Cassell, and Bhatnagar 2021).

Respondents noted complexities of sharing data from the unfamiliar process of wastewater monitoring for SARS-CoV-2. Many mentioned the benefits of transparency (immediately and publicly sharing wastewater results), such as building trust and encouraging protective behaviors. Alternately, several respondents expressed concerns that public access to results could incite unnecessary panic or cause people to second-guess the college’s responses. One respondent noted positive feedback from parents of students who observed with gratitude that the institution was taking a proactive step to maintain students’ health by monitoring wastewater. Another noted they refrained from publicizing dormitory wastewater results in order to avoid creating a stigma against students from a particular demographic or interest group who resided in “themed dorms.” Colleges made different tradeoffs between transparency, sensitivity, and privacy depending on their campus culture, leadership, and confidence in wastewater results.

### Key elements of success and ongoing challenges

Respondents offered several insights into the key elements that contributed to progress in wastewater surveillance at their colleges as well as ongoing challenges. Common themes are summarized below.

Respondents identified a wide range of elements of the wastewater monitoring process that worked well (self-defined “successes”), ranging from technical to educational to social.

- **Collaboration:** Nearly all respondents praised cooperation among faculty, facilities staff, university administration, and, in several cases, the staff of wastewater treatment facilities. One faculty member noted the wastewater monitoring effort had led to “amazing collaborations and research opportunities that normally don’t fall in my scope of work.” Respondents also reported new partnerships with other colleges, community leaders, and government agencies (e.g., public works and public health). Several noted that communicating with practitioners in other colleges helped them create successful workflows. One respondent noted that “there has been an *incredibly* collegial attitude about wastewater testing during the pandemic; it’s like nothing I’ve ever seen before!”
- **Student engagement:** Several of the colleges that engaged students in sampling and analysis highlighted students’ enthusiasm and learning experiences as a benefit. One student who was involved in her college’s wastewater surveillance noted an “immense feeling of pride and satisfaction…The knowledge and skill set I have developed are so valuable, and the work we did will make such a difference for our community and the environment!” (Siena College 2020) Others noted that students contributed insights about campus behaviors (e.g., location of parties) that informed wastewater sampling locations and helped spread the word to others about the value of the wastewater monitoring program.
- **Motivated staff:** Many respondents praised the involvement of “amazing” staff. One respondent suggested that it was most productive to find the people on campus who were “eager and willing” to help with wastewater monitoring, whatever their role, and work with them to collect samples.
- **Support from college administration:** Administrators who supported college wastewater monitoring efforts with resources – including financial support, staff time, and release from teaching obligations – were vital. Several administrators adapted the college’s policies to address urgent needs and streamline slow-moving bureaucratic processes. As one respondent reflected, “universities are not flexible enough to handle the rapid and nimble responses required to address a pandemic (e.g. hiring and purchasing processes), so you need to have the president’s support to help bend rules and find work-arounds to get things done.” Respondents also noted that high-level support was helpful because the steep learning curve of wastewater surveillance often resulted in unexpected challenges, delays, and costs. As one respondent said, “Be prepared to pay overtime.”
- **Problem-solving and adaptation:** Finding resourceful solutions to local challenges was a hallmark of many of the college wastewater monitoring efforts. Respondents reported adapting to changing student population sizes and living situations, creatively sampling from less-accessible sewers, and developing inventive work-arounds to supply-chain disruptions. In addition, many respondents noted they improved analytical methods in the laboratory to gain greater sensitivity, reduce turn-around time for results, and reduce costs. Many local solutions were made possible by support from collaborative networks with practitioners from other colleges and wastewater agencies.

Respondents also cited many challenges, most of which related either to aspects of wastewater surveillance or complexities of interpreting results. Some of the most commonly cited technical challenges included:

- **Supply chain delays:** Respondents noted delays resulting from limited availability of autosampler pumps, centrifuge equipment, and RNA extraction kits.
- **Obtaining representative samples:** Even with composite sampling, obtaining representative samples at the dormitory scale is challenging due to issues with non-homogenous wastewater (variable fecal concentration), low-flow conditions, and autosampler intake clogging.
- **Collection system logistics:** The ease of wastewater sampling depends on the college’s physical layout. For example, mapping sewer pipes and installing autosamplers in the plumbing of older campuses may be more logistically challenging due to the age and complexity of their sewer systems. Other colleges had to obtain special permits to lift manhole covers in city streets to obtain wastewater samples, figure out how to safely enter a confined sewer space, and protect autosamplers from theft or vandalism.
- **Developing laboratory methods:** Many college laboratories faced challenges developing appropriate techniques for concentration, extraction, and data analysis. As one respondent noted, “There are so many little lessons learned from making mistakes…there is going to be trial and error.”
- **Safety protocols:** Researchers are still unsure how persistent infective SARS-CoV-2 is in wastewater (Amoah, Kumari, and Bux 2020), leading to uncertainty about the necessary levels of laboratory disinfection, equipment cleaning, and protective equipment required by personnel to collect and analyze samples. Biosafety protocols posed a hurdle for many college laboratories.
- **Timing:** In order to effectively inform decisions (e.g. follow-up testing of students), wastewater monitoring results need to be available quickly. Although sample processing time was typically under 12 hours, sample collection, lab workflows, shipping, and staffing limitations often delayed availability of results.
- **Scaling up from research to production:** Many colleges initiated wastewater surveillance through pilot-scale research projects. The complexity of expanding to campus-wide monitoring was frequently underestimated. Associated challenges included human resources, training, biosafety regulations, supplies, equipment, and space as they scaled up their efforts.

In addition to these technical challenges, many respondents noted the complexities involved in interpretation, communication, and use of wastewater results. There is currently no standard guidance for interpretation of wastewater results. In particular, several colleges noted difficulty “reconciling results from the wastewater with individual testing.” One respondent found it difficult to explain to administrators why SARS-CoV-2 RNA was not detected in wastewater results from the “isolation dorm” that housed students known to be infected with COVID-19. Similarly, several colleges reported multiple instances of detecting the virus RNA in dorm wastewater, testing residents, and finding no positive individual test results. There are multiple possible explanations, including false negatives during clinical testing, low compliance with clinical testing directives, fecal contributions by non-residents, and convalescent students back in residence. However, it was challenging to explain these possibilities to anxious students, administrators, and members of the public. These examples highlight the complexities of communicating uncertain but highly salient information in real time. As one respondent said, “One must interpret the data the best that one can and not overstate.”

Participants gained experience interpreting trends in their particular setting over time. One respondent noted, “After a while we learned that three points with a clear positive slope meant there was an increase in cases in a dorm (an outbreak), but…did not accurately predict the <u>number</u> of people we would find.” Many colleges struggled with effective messaging to diverse audiences. For example, one noted that the “gross factor” associated with their initial choice of terminology distracted students from their public health message. Another noted that emails recommending testing following positive wastewater results were so frequent that students became inured, resulting in reduced compliance with follow-up testing.

Despite such challenges, many respondents remained positive about the potential for wastewater surveillance to enhance colleges’ pandemic responses. Several respondents shared personal reflections about their ongoing support for wastewater monitoring.

- “There is a huge amount of value in getting negative results out of dorms, and that is underestimated. Every time I get a zero [no detection of SARS-CoV-2 RNA from wastewater] that is a comfort. There could be a case there that wasn’t caught, but there’s a very low probability that an actual outbreak is occurring.”
- “The director of our regional health department said our college’s wastewater monitoring has ‘protected our community from wider spread infections.’”
- “Wastewater testing allows use of limited testing resources to maximum benefit.”
- “We successfully stopped an outbreak based on this surveillance.”
- “Wastewater testing gave us a short, advanced warning of our outbreak, enough to mobilize mass testing and request additional resources. It probably gained us a few days in identifying and isolating students.”
- “The benefit to cost ratio is huge.”
- “Wastewater is a major piece of the puzzle in preventing outbreaks in the dorms…I truly believe the tremendous efforts of the individuals who have worked on these projects controlled outbreaks, kept campuses open, and most likely saved lives.”

One caveat was that wastewater surveillance adds more value for some colleges than others, and that some “universities with extensive clinical testing are hesitant to utilize wastewater” because they do not think it adds valuable information. Others see the two approaches as complementary, with wastewater providing early warning and a check on untested individuals.

Overall, respondents painted a picture of developing wastewater surveillance as a collaborative learning process involving diverse on- and off-campus stakeholders. Particularly at colleges where the surveillance efforts originated with research, these efforts often pulled faculty into unfamiliar roles. Several faculty members who initiated wastewater monitoring provided specific advice for others in similar positions (see Supplemental Material: Insights and advice to fellow faculty engaging in wastewater monitoring).

## A process-oriented framework for wastewater surveillance

These findings suggest that there is no single ideal, universally applicable approach to wastewater monitoring on college campuses. Rather, each campus experienced an iterative, user-informed process that involved identification of unique information needs, sewage infrastructure, opportunities for wastewater sampling and analysis, ways to interpret results for decision makers and approaches to communication (Figure 2). The interplay between these different factors informed development of each college’s wastewater surveillance strategy. Although the idea of a “playbook” for wastewater monitoring is appealing, the diversity of these 25 colleges’ experiences suggests that it may be more appropriate to design wastewater surveillance as a process of collaborative learning and adaptation. Based on these 25 colleges’ experiences, we developed a framework for structuring such an iterative process. This process is delineated in Table 3 through a series of questions for consideration at each step.

**Table 3:** A framework for designing a campus wastewater monitoring system

| **Framework element** | **Key question** | **Factors for consideration** |
| --- | --- | --- |
| Information needs | Who will use the information? What information do those users need from wastewater? | • Target individual testing and contact-tracing resources<br>• Identify SARS-CoV-2 trends over time<br>• Compare on- and off-campus trends<br>• Limitations/uncertainties of results<br>• Resources available (expected value of sample information) |
| Wastewater infrastructure | How can the sewage infrastructure be accessed? | • Identify and create maps of sewer system<br>• Assess accessibility of sampling sites<br>• Coordinate with municipal wastewater agency and/or campus facilities staff |
| Sampling plan | How can we sample wastewater? | • Select sampling locations<br>• Consider tradeoffs between composite samples, passive samples, or grab samples<br>• Determine sampling frequency (samples/week)<br>• Identify who will collect samples |
| Wastewater analysis | Who can analyze and interpret wastewater samples? | • Assess wastewater testing options (on-campus or private lab, cost, turnaround time, capacity, safety regulations, etc.) |
| Data Interpretation and use | How can findings inform decisions? | • Determine who will interpret data and assess trends<br>• Access public health information needed to contextualize data (e.g. number of people in quarantine or recently recovered)<br>• Consider range of decision outcomes (e.g. testing, messaging, limiting gatherings, remote instruction) |
| Communication plan | What is the most effective way to share findings with appropriate audiences? | • Who should be involved in messaging?<br>• Who are the key target audiences?<br>• How can messages best be communicated to intended audiences (e.g. signs, email, social media, website, etc.)? |

**Figure 2.**
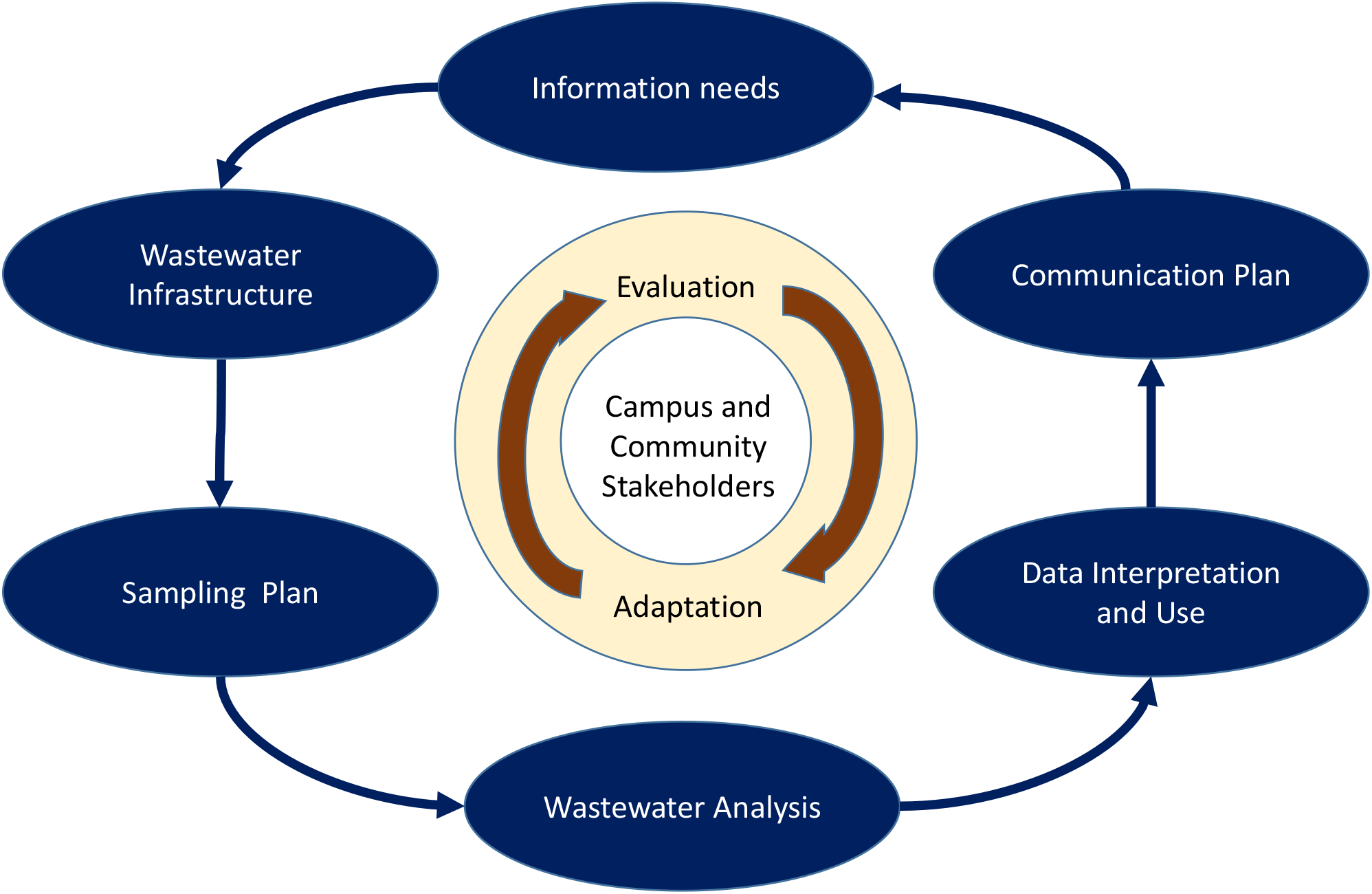
An iterative, process-oriented framework for wastewater surveillance at colleges.

### Information needs

Different institutions have different information needs, ranging from saving money by minimizing the need for clinical testing, to identifying the presence of infected individuals who were not tested, to reassuring the local community that students are not spreading COVID-19. The potential for wastewater surveillance to meet these needs is shaped by many factors, including the proportion of students who reside on campus and the current community-wide infection rate. It is important to involve potential information users, including administrators, student life, communications, and community stakeholders in designing the wastewater surveillance system. Sources and implications of uncertainty, alternative approaches, and resource requirements are a key part of this discussion. Clarifying the expected use of wastewater results can guide subsequent decisions about the sampling plan and tradeoffs when resources are limited.

### Wastewater infrastructure

Understanding the local sewer system is essential, since physical layout and accessibility often constrain the wastewater monitoring approaches. Many colleges lack an accurate map of their wastewater infrastructure, particularly as it interfaces with the surrounding community. The next step is to identify potential sampling locations (manholes, building cleanouts, wastewater treatment plants). Sewer systems are designed for efficient wastewater conveyance, not for public health surveillance – so collecting samples at the best locations for informing public health decisions may not be possible. This can complicate colleges’ efforts to use wastewater testing to identify specific groups of students for testing. For example, single dormitories may have multiple sewer outlets, share outflows with adjacent dorms or public buildings (e.g. dining halls), or be integrated with community systems. The physical layout of wastewater infrastructure must be considered in the development of the sampling strategy, the interpretation of results, and determination of follow-up actions.

### Sampling plan

Once the flow of wastewater is understood, a sampling plan can be designed to meet identified information needs. Choosing sampling sites requires input from decision makers, facilities staff, and other stakeholders to assess the merits of various options. For example, certain sites may be difficult to access (either physically or legally, as with manholes located in public streets), have potential for clogging, have inadequate flow, pose a risk to security of autosamplers, or be vulnerable to extreme weather conditions. The choice of autosamplers, passive samplers (absorbent swabs), or grab samples should include consideration of equipment costs, staff time, wastewater heterogeneity, and institutional characteristics. For example, since peak flows are not as predictable when students do not have to leave their housing at a specific time to attend class, composite sampling may be especially useful at schools with virtual instruction. However, composite samples can also dilute SARS-CoV-2 signals, particularly in low prevalence areas. Passive samples may be cost-effective, but there have not yet been robust studies comparing results between composite samplers and passive swabs. Finally, sampling frequency must be determined. This may involve tradeoffs between resources (costs, staff time, etc.) and ability to rapidly identify trends in the data. Decisions about who will collect samples depend on multiple factors including cost, safety regulations, and timing of analysis. Colleges may establish different sampling schemes for different locations: for example, by using student labor in accessible on-campus locations and employing contractors for off-campus sample collection, or by varying frequency of sampling at different sites.

### Wastewater analysis options

For colleges with on-campus laboratories capable of performing wastewater analysis, the choice of analysis approach may be straightforward. However, on-campus labs will need to plan carefully to scale up their capacity. For colleges relying on commercial analysis services, considerations include cost, turnaround time, and reliability. The total time for shipping, analysis, and return of results may vary significantly among commercial labs, and rapid return of results is essential to end users. The recent proliferation of commercial laboratory services means that it may be difficult for colleges to identify differences in limits of sensitivity, reliability, reporting formats, and quality control.

### Data interpretation and use

Despite the desire expressed by many colleges to have a predetermined “end use protocol” for wastewater results, contextual information and expert human judgement in interpreting results are critical. Wastewater monitoring results are most informative when integrated with individual testing data and other contextual information about sample representativeness, the results of laboratory positive and negative controls, the boundaries of sewershed catchment areas, and the number of infected and recovering individuals in each catchment. Use of wastewater data depends upon the college’s unique social and institutional dynamics. For example, the ability to follow up on a positive wastewater signal with individual diagnostic testing may be determined by whether the college is able to mandate student testing, whether students tend comply with or evade testing requirements, and whether students are willing and able to self-isolate. Different institutions have varied potential public health interventions depending on their resources, physical structure, student body size, and other constraints (e.g. public versus private). This breadth of considerations suggests that a team of individuals with diverse experience is needed to interpret results on an ongoing basis, ideally including expertise in environmental engineering, epidemiology, biostatistics, facilities management, campus operations, student life, and communications.

### Communication plan

It is important to prepare a communication plan prior to detecting spikes in wastewater. Communication plans should engage a wide range of stakeholders, including wastewater experts, university communications, legal experts, and student life professionals. Students may also inform effective messages and communication approaches. Each college should identify appropriate visualization tools for its intended audiences. Examples include using color to highlight data trends in particular locations; superimposing data on a map of campus residence halls; or showing trends in wastewater data along with trends in clinical cases. Finally, the communication plan should carefully consider the advantages and disadvantages of transparency about wastewater data given inherent uncertainties, privacy considerations, and contextual factors.

### Evaluation and adaptation

As the cyclical design of Figure 2 indicates, experience and changing circumstances may require adaptation of initial wastewater surveillance plans. Colleges should establish structures, metrics, and collaborative processes for ongoing evaluation and adaptation. Most fundamentally, it is important to revisit whether the initially identified information needs are being met, and if not, whether the wastewater surveillance program can be adjusted to do so. Additional resource needs may be identified. Alternately, expectations about how wastewater results can support the college’s COVID-19 management efforts may need to be altered.

## Discussion

This analysis is limited by the information provided by the 25 colleges that chose to participate in this study. This small pool may not be representative of the many institutions that have implemented wastewater surveillance. In most cases, the information provided represents the knowledge of one key informant at each college. Future in-depth case studies could shed light on varied perspectives by multiple stakeholders at each institution. Nonetheless, the wide range of approaches taken by these cases provides key insights to better understand the potential for wastewater monitoring to inform colleges public health decision making.

This study highlighted some of the research needs related to wastewater monitoring for SARS-CoV-2 on college campuses. While several respondents stated unequivocally that their wastewater monitoring programs were worth the effort, others voiced the need for a more systematic assessment of the costs and public health benefits of wastewater monitoring at colleges. More research is needed to determine how wastewater surveillance and individual clinical testing for SARS-CoV-2 can be most effectively paired to reduce COVID-19 transmission. There is also an urgent need for better understanding of how colleges’ varied social and decision-making contexts (i.e., privacy concerns, consent, communication, baseline health of the populations, and degree of administrative controls over the social environment) affect their wastewater surveillance efforts. For example, wastewater monitoring may be particularly useful in the setting of public universities, which may be less able to compel students’ compliance with clinical testing.

Research to assess the sensitivity of low-cost sampling methods is needed. Comparisons of results from grab, passive swab, and composite samples at the building scale could help resource-limited institutions make appropriate choices. Ultimately, a clear understanding of the sensitivity of each of these approaches for detecting infected individuals in a building would be very helpful. Additional research to understand better the variability associated with wastewater data is critical to its effective use.

Many respondents expressed a need for protocols for communication and use of wastewater results. To help inform such guidelines, social science research is needed to help identify effective ways to communicate uncertain results from wastewater surveillance, to motivate behavior, and to support decisions using multiple sources of information. An in-depth analysis of the ways in which different colleges (and other residential facilities) have interpreted and communicated results of wastewater monitoring, along with corresponding changes in behavior and case rates could elucidate some of these key information needs.

The diversity in approaches across the colleges included in this study was largely driven by differences in physical infrastructure layout, research expertise, financial resources, institutional characteristics, and leadership support. Research that informs cost-effective implementation of wastewater monitoring at institutions with limited technical, financial, and human resources is essential to promote equity in both health and educational outcomes.

## Conclusions

This initial overview of wastewater monitoring at colleges across the U.S. reflects a wide variety of experiences. These efforts were started by different stakeholders (faculty, staff, administration, public health officials) for different reasons. Their diverse goals, combined with varied funding, physical conditions, research expertise, and technical capacity, resulted in approaches that vary in nearly every dimension (e.g. number and types of sites sampled, frequency and methodology of sampling, analysis methodology, and use of data in decision-making).

Despite differences in their approaches, common themes emerged from these colleges’ experiences. Most colleges encountered unexpected challenges in the design and implementation of wastewater surveillance, resulting in rapid learning and frequent recalibration of expectations. The vast majority faced challenges in how to interpret, communicate and use wastewater results to inform their pandemic response. Collaboration – both within and outside of the institution – was reported as essential to success in nearly every case.

These initial experiences provide many lessons, both for other colleges contemplating implementing wastewater monitoring as part of their broader COVID-19 surveillance systems, as well as for other types of institutions and community-level monitoring efforts. These lessons include the need for a systematic assessment of wastewater infrastructure, sampling options, and consideration of data use when designing the system. In addition, these experiences indicate that developing and implementing effective wastewater surveillance programs at colleges requires a collaborative multidisciplinary process, in which diverse campus and community stakeholders iteratively evaluate and adapt their strategy to best inform public health action.

## Data Availability

The interview transcripts used for the research contain identifying information and are not available to the public.

## Acknowledgements

The authors would like to thank these individuals for their important contributions to this research and information about their colleges’ wastewater surveillance systems: Aaron Best, Walter Betancourt, Cindi K. Brinkman, Lifang Chiang, Erik R. Coats, David Freedman, Elmer Johnson, Rob Knight, Karl Korfmacher, Erin K. Lipp, Erin A. Mack, Christopher Maroney, Simona Matsoyan, Ian Pepper, Shalina Shahin, Lachlan Squair, Eva M. Top, and Rogelio Zuniga-Montanez. Dr. Korfmacher’s work on this project was supported in part by the National Institute of Environmental Health Sciences grant P30 ES001247. Dr. Harris-Lovett’s work on this project was supported in part by the Catena Foundation. The content of this manuscript is solely the responsibility of the authors and does not necessarily represent the views of contributors, interviewees, or institutions included in the study.

