## Supplemental Text 1. for "Wastewater surveillance for SARS-CoV-2 on college campuses: Initial efforts, lessons learned and research needs"

**Text S1. Insights and advice for fellow faculty engaging in wastewater monitoring**

“I am not a doctor and I’m not out there on the front lines...but this is one thing I can do to help my community.”

“This has become very stressful each week to get data out when we intend while ensuring that the quality control checks out. People are now looking for weekly updates. It's nerve wracking compared to the traditional way that we collect data and publish.”

[Wastewater monitoring] “requires staffing that is dedicated; think of it as a production-level project -- not an academic one -- you need data that is good enough and that can be used for decision making.”

“It is a lot of work to put together a high frequency, multi-site monitoring system -- definitely communicate; don't reinvent any wheels; leverage existing expertise that you may not realize you have; collaborate with local institutions.”

“As a scientist, when I suggested wastewater surveillance, I didn't realize how challenging this project would be from the Emergency Management side of things.  If you bring wastewater to the table, you might be asked to wear many hats to get the program operational.”

“Sewer surveillance is not for the faint of heart.  It requires making decisions on limited data, learning on the fly what the strengths and limitations of the technique are, learning how to communicate with nervous administrators, elected officials and general public, and being willing to take a full-time job on the top of your regular full-time job.  That said, it has also been one of the most exciting and rewarding experiences of my career. The level of collaboration both within the university, across disciplines and with facilities and housing, as well as the city, county and state has been nothing short of phenomenal and amazing.”

“This epidemic has built bridges between people that have never met.”

“The best thing has been the good feeling that I have about working on something that may benefit the health of people in my community.”

“Establishing a team with the appropriate skill set and expertise is key.”
